# Exploring factors affected timely receiving intravitreal anti-VEGF treatment in patients with Diabetic Macular Edema: a qualitative interview study analyzed with COM-B model

**DOI:** 10.1101/2024.04.09.24305579

**Authors:** Shu Li, Jiani Pan, Yan Xu, Shiyu Tian, Zhengyue Dai, Qiong Fang

## Abstract

**Objectives:** To explore factors affected untimely receiving intravitreal anti-Vascular endothelial growth factor (VEGF) injection based on the Capability, Opportunity and Motivation-Behavior (COM-B) model in patients with Diabetic Macular Edema (DME) and regard these factors as main targets for interventions.

**Design:** An exploratory qualitative study was conducted using semi-structured interviews with patients with DME. The COM-B model was used to guide data collection and analysis.

**Setting:** The study was carried out in outpatient department of ophthalmology in China.

**Participants:** 24 patients with DME who experienced anti-VEGF treatment at least one injection within one year were recruited using convenience and purposed sampling.

**Results:** The themes and subthemes were identified. Physical capability included (1) lack of knowledge, (2) high treatment expenditure, and psychological capability included prioritized glycemic control rather than anti-VEGF. Social opportunity included (1) no anti-VEGF available, (2) Recommended eyedrops, laser and oral drug but not mentioned intravitreal anti-VEGF injection, (3) As an optional therapy, selected more convenient treatment rather than anti-VEGF agents, and physical opportunity included (1) no confidence in treatment from doctor, (2) communication between doctors and patients. Reflective motivation included (1) outcome expectancies, and automatic motivation included (1) fear of injection, (2) fear of blindness.

**Conclusion:** We should attach importance to these eleven factors, especially to effective interaction between doctors and patients, and doctor’s authoritative treatment advice, which interventions were based on in the future.

**Strengths and limitations of this study:**

⇒ The qualitative design was used to understand factors affecting timely intravitreal anti-VEGF injections in patients with DME and to explore the potential measures to change them.
⇒The study highlighted the importance of efficient communication between clinicians and patients.
⇒The study didn’t consider other clinical variables, such as the severity and duration of diabetes, which should be included in the future studies.

## Introduction

Diabetic macular edema (DME), a manifestation of diabetic retinopathy, that impairs central vision and has a profoundly detrimental impact on vision-specific function, mobility, independence and quality of life (QoL).[1]

Vascular endothelial growth factor (VEGF) is an important mediator of abnormal vascular permeability in DME. Intravitreal injections of anti-VEGF agents have been shown to be superior to laser photocoagulation of the macula, and become first-line therapy in patients with DME in the past 15 years.[2,3] The treatment regimen of DME is composed of three continuous monthly anti-VEGF injections followed by additional injections when necessary according to OCT results and visual acuity improvement during the first year of treatment.[2] However, outcomes observed in clinical practice do not reach those in clinical trials,[4] which due to far less injection frequencies in clinical practice than those in clinical trials, in other words, failure to receive intravitreal injection as prescribed and keep scheduled follow-up reduces therapeutic effect and results in poorer outcomes.[5]

Previous researches on adherence to anti-VEGF treatment mainly focused on the foreign population.[3,5–7] There is a paucity of literature on Chinese patients, therefore, it is necessary to understand factors affected timely receiving intravitreal anti-VEGF treatment in China.

Some researchers and organizations suggest that intervention developed on theoretical models are more likely to be effective[8,9] and are tailored to the population and context in which the target behaviors are implemented.[10] One such approach is the Behavior Change Wheel(BCW), at the hub of it, the COM-B model aims to facilitate a behavioral diagnosis.[10] The COM-B model also been used in conjunction with the Theoretical Domains Framework (TDF) to better understand the impact on the target behavior.[9–11] Therefore, the aim of this study was to identify factors affected timely receiving intravitreal anti-VEGF injection in patients with DME based on COM-B model, and to provide the basis for interventions to improve adherence to anti-VEGF treatment.

## 1. Method

### 1.1. Design and setting

A larger, three-phased mixed methods study is being conducted to develop a theory-based intervention to improve adherence to anti-VEGF treatment in patients with DME. This present study was Phase 1 to use a theoretical framework to identify the factors that influence intravitreal injection of anti-VEGF agents. We used a qualitative descriptive design to conduct semi-structured interviews with patients undergoing anti-VEGF treatment. The research was performed consistently with a reporting framework of Consolidated Criteria for Reporting Qualitative Research.

Ethical approval for the study protocol was obtained from the ethics committees of Shanghai General Hospital, Shanghai Jiao Tong University School of Medicine (No. 2022-KY-021).

### 1.2. Recruitment of participants

Convenience sampling was used to recruit the participants from seven provinces in China between February and May 2022 to represent different patient demographics and access to anti-VEGF treatment. The eligibility criteria for participants were as follows: (1) aged ≥ 18 years; (2) diagnosis of DME and at least one prior intravitreal injection of anti-VEGF agents; (3) the ability to communicate in Chinese. We selected patients of different ages, literacy levels, sexes, and diagnoses to ensure a broad selection.

### 1.3. Data collection

The study team developed an interview guide to better understand patient’s perspectives on anti-VEGF treatment based on the TDF (Table 1). Before the interview, participants were informed of the purpose of the study and the content of the interview and signed written informed consent.

**Table 1.**
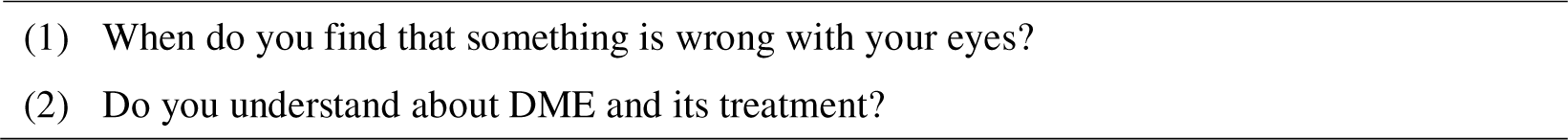
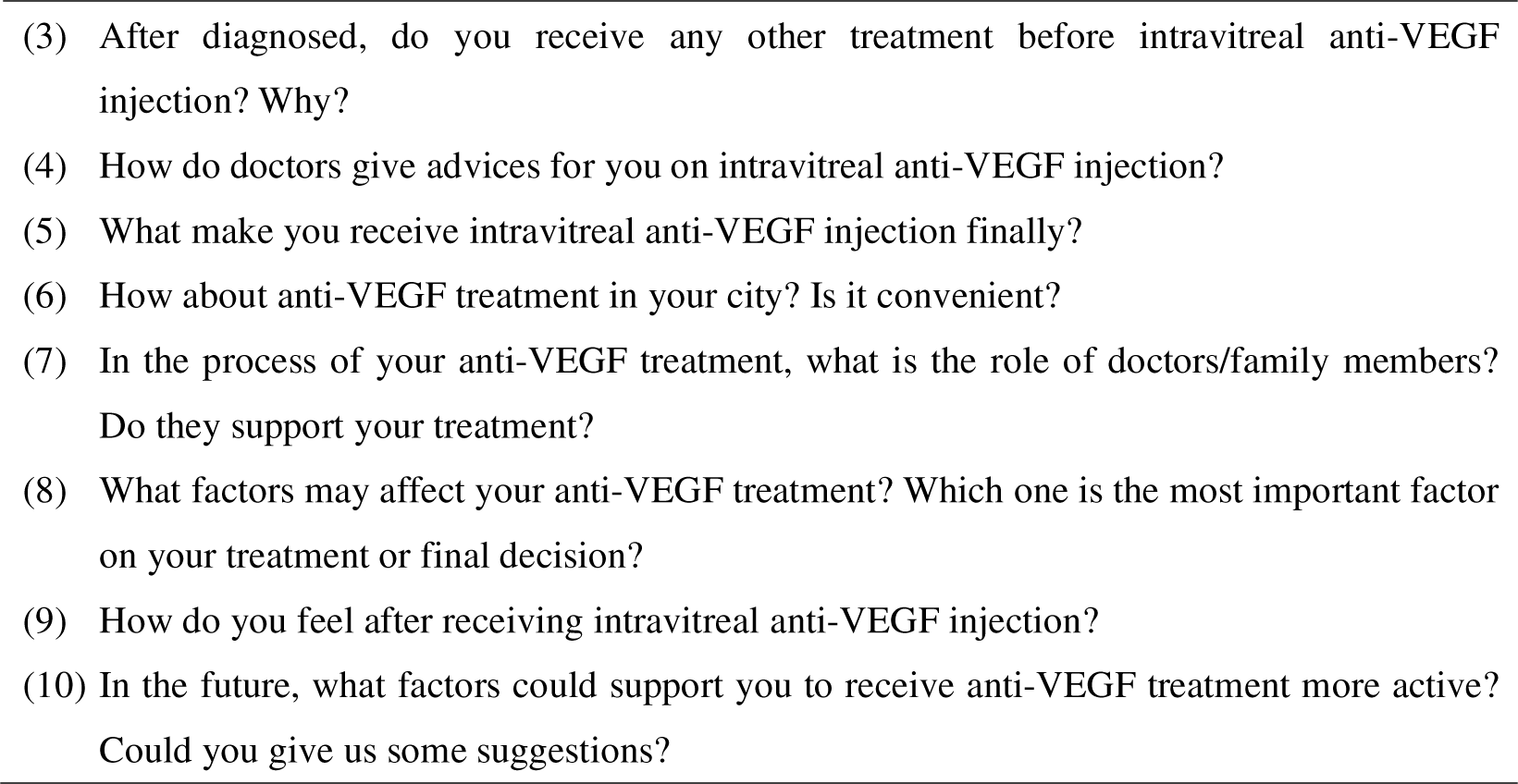
Interview questions

Semi-structured interviews by face-to-face or telephone were conducted by an experienced qualitative researcher (LS) who received initial and ongoing qualitative training from Shanghai Jiao Tong University School of Nursing and by another senior qualitative researcher (XY) and were audio-recorded and transcribed verbatim within 24 hours. The transcripts were given to the participants for any corrections or comments to clarify unclear answers. We also conducted a brief demographic survey at the beginning of each interview.

### 1.4. Data Analysis

Participants were numbered according to the interview order (Participant 1, Participant 2…). The transcripts were coded independently by two researchers (PJN, TSY). Then the thematic analysis was conducted to analyze the data by Colaizzi, following (1) read all the interview materials carefully, (2) extracted important statements in line with the study phenomenon, (3) encoded recurring important statements, (4) gathered the encoded statements, (5) wrote a detailed and complete description, (6) identified the similar statements and (7) returned to the participants to verify the authenticity of the content. The research team constantly compared the analysis and results to ensure accuracy.

## 2. Results

### 2.1. Participant characteristics

The study engaged 24 participants (Table 2), with city coverage Tier 1-3, Tier 4 and county city. Mean interview time was 45min (range 35-75min).

**Table 2.** Demographic characteristics of participants who experienced anti-VEGF therapy

| Characteristic | N (%) |
| --- | --- |
| **Total Number** | 24 |
| **Mean Age (range)** | 60.4 (28-68) |
| **Gender** |  |
| Male | 17(70.8) |
| Female | 7 (29.2) |
| **Marital status** |  |
| Married | 18(75.0) |
| Unmarried | 1 (4.2) |
| Divorce/Widowed | 4 (15.8) |
| **Education** |  |
| Primary school and below | 2 (8.3) |
| Junior high school | 17 (70.8) |
| Senior high school | 3 (12.5) |
| College or university and above | 2 (8.4) |
| **Occupation** |  |
| Employed | 5 (20.8) |
| Retired | 19(79.2) |
| **Cities** |  |
| Urban | 13 (54.2) |
| Suburban | 11 (45.8) |

### 2.2. Summary of themes and subthemes

Eleven subthemes emerged from the COM-B domains are shown in Table 3.

**Table 3.**
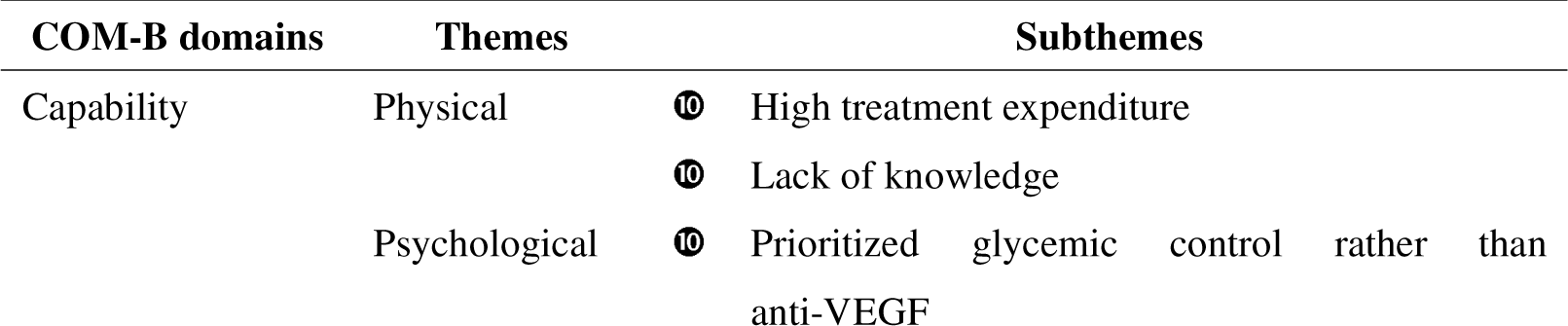
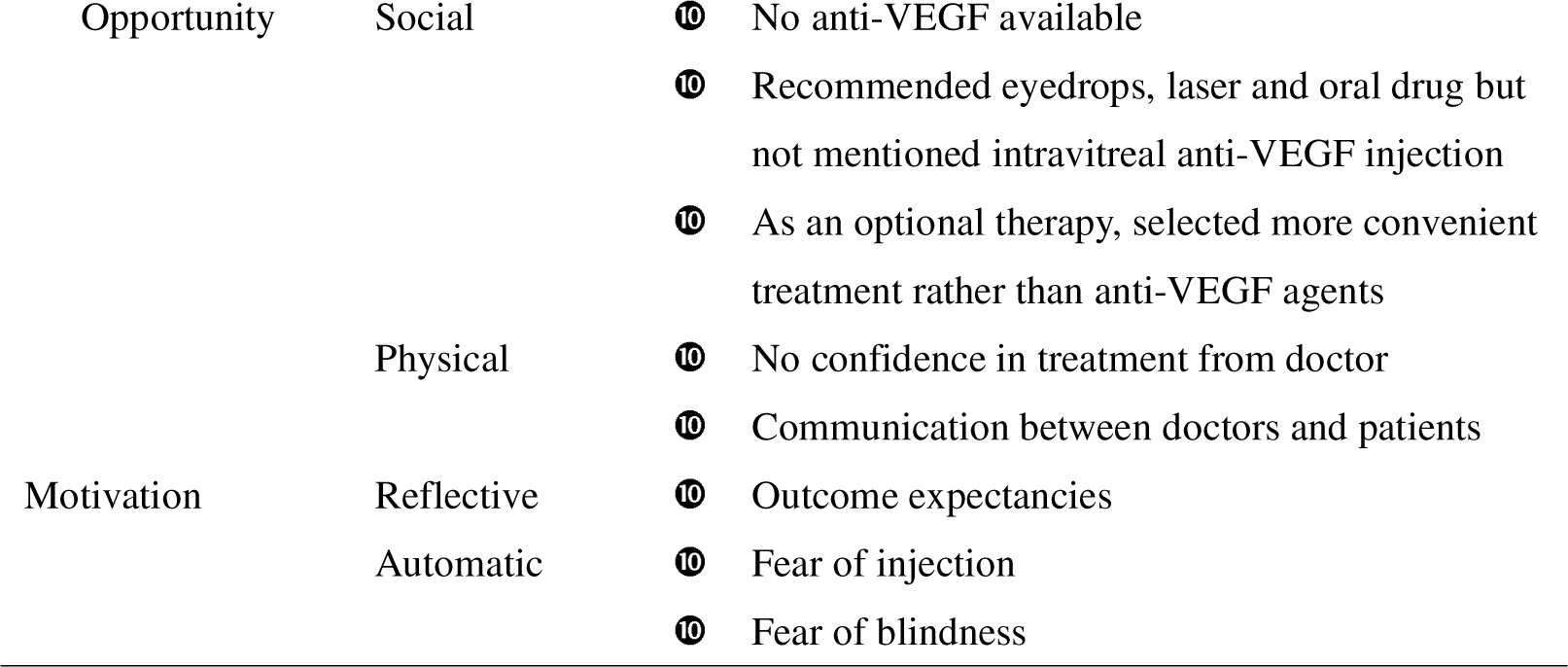
Twelve subthemes emerged from the COM-B domains

#### 2.2.1. Capability

##### 2.2.1.1. Physical capability

One subtheme ‘high treatment expenditure’ was identified in terms of physical capability, which delayed the decision on anti-VEGF treatment.

###### High treatment expenditure

Some participants claimed that anti-VEGF treatment was too expensive to inject for 2-3 doses. When their visual impairment severely affected their daily lives, they finally received intravitreal injection.

> *When I asked doctor about the total cost of treatment, he said ¥ 6,000 per one injection and could not be reimbursed. How could I afford that? It was too expensive, so I gave up…* (Participant 6)
>
> *My eyesight was blurred for many years, three years ago, I was diagnosed with DME, doctor told me that I should receive anti-VEGF injection, due to high cost of treatment, I gave up. Now my daughter is working, my economic condition has significantly improved, so I want to receive anti-VEGF treatment…* (Participant 1)
>
> *My right eye missed the best time of treatment and there was no value of intravitreal injection anti-VEGF agents, so I received anti-VEGF treatment only in my left eye, but it was too expensive that I can’t afford it, thus, I only finished one injection and went back to my hometown where I could use my medical insurance…* (Participant 24)

##### 2.2.1.2. Psychological capability

Two subthemes were identified concerning psychological capability: (1) lack of knowledge, (2) prioritized glycemic control rather than anti-VEGF.

###### Lack of knowledge

Some participants thought some symptoms as visual deterioration that were the normal process of aging or ‘with cataract’.

> *I can’t see the glasses on the table clearly, I think it is my eyes are aging, then I go to an optical shop and selected presbyopia glasses there…* (Participant 7)
>
> *I found the wall was twisted, I thought I was too tired, so I didn’t care about it…* (Participant 23)
>
> *In the beginning, I just found the wall was curved, I thought I had cataract and paid little attention on it…* (Participant 2)

###### Prioritized glycemic control rather than anti-VEGF

Some participants prioritized glycemic control rather than anti-VEGF treatment, and counted on glycemic control to alleviate symptoms in the eyes.

> *Doctor in a county hospital told me the most crucial thing is sugar control, so I I treated my diabetes immediately. But after controlling glycemic well, there was no improvement in my vision acuity, even worse than before, thus, I had to go to an ophthalmology hospital and was diagnosed with DME and receive anti-VEGF treatment there…* (Participant 17)

#### 2.2.2. Opportunity

##### 2.2.2.1. Social opportunity

‘No anti-VEGF available’, ‘Recommended eyedrops, laser and oral drug but not mentioned intravitreal anti-VEGF injection’ and ‘As an optional therapy, selected more convenient treatment rather than anti-VEGF agents’ were the subthemes which identified and presented in terms of social opportunity. Some participants narrated that it was very difficult to find a hospital where they can get appropriate treatment in their hometown especially in suburban cities. Therefore, it is common to visit multiple hospital to receive anti-VEGF treatment due to low trust toward local hospitals.

###### No anti-VEGF available

Some participants expressed that there were no anti-VEGF agents in their hometown, they had to transfer to other hospitals in provincial capitals or large cities.

> *I went to the hospital because of pain in my right eye. After finished some examination, I was diagnosed with DME, but anti-VEGF agents is not available in that hospital. Doctor advised me I should go to large hospital and receive intravitreal injection anti-VEGF agents there, but COVID-19 outbreak delayed treatment…* (Participant 4)

###### Recommended eyedrops, laser and oral drug but not mentioned intravitreal anti-VEGF injection

Some participants narrated that they were diagnosed with DME and doctors recommended eyedrops, laser and oral drug but not mentioned intravitreal anti-VEGF injection.

> *When I read the newspaper, the writing blurred and danced before my eyes at first, I believed it was the process of aging and paid little attention on it, six months later, I found that my vision acuity was becoming worse, so I went to the hospital and doctor told me my eye was diagnosed with DME, but nothing more to tell, then I had to went to another hospital…* (Participant 2)
>
> *My initial symptoms were not serious just with eyes redness and discomfort, so I didn’t consider it as a severe disease. I went to nearest hospitals with my wife and doctor told me that I could treat with eyedrop first to relieve my symptoms…* (Participant 10)
>
> *When I found something wrong with my eyes, I went to nearest hospitals where my family members treated their general disease and doctor told me that I could treat with eyedrop first to relieve my symptoms, you know, there is not much choice for me in my hometown…* (Participant 12)

###### As an optional therapy, selected more convenient treatment rather than anti-VEGF agents

Some participants narrated that doctors mentioned intravitreal anti-VEGF injection but not provide much information. When knew it for the first time, they were uncertain about therapeutic effects, and as an optional therapy, tended to choose more convenient treatment as their first choice, such as laser, eyedrops and oral medications.

> *I felt blurred in my eyes, so I went to local hospital, after many examinations, I was diagnosed with DME, while I first knew I had diabetes. I went to endocrine department, doctor told me that I should control my glycemic first. I also went to ophthalmology department in the same hospital and doctor told me anti-VEGF treatment as an optional therapy, I can treat with eyedrop first…* (Participant 8)
>
> *I felt wall was twisted but I didn’t care about it. I went to a local hospital until black spot appeared all the time in my eyes. Doctor told me there were something wrong with my macular, after examining, I was diagnosed with DME. Doctor said it can treat with laser, eyedrop and intravitreal injection of anti-VEGF agents, so I selected eyedrop first…* (Participant 23)

##### 2.2.2.2. Physical opportunity

###### No confidence in treatment from doctor

Doctor suggested trying intravitreal anti-VEGF injection, and the therapeutic effect was not clear.

> *When I finished ophthalmologic examinations and was diagnosed with DME, doctor told me that I can try receiving 2∼3 injection of anti-VEGF agents which may be effective of my eyes…* (Participant 13)
>
> *Doctor told us that anti-VEGF treatment may showed minimal response, with some patients sensitive and others not, but it is a worthwhile method to try, so he asked us to consider it clearly. Finally, I tried it…* (Participant 16)

###### Communication between doctors and patients

Sometimes, especially when doctors were unfamiliar with, they were afraid of rejection which hindered them from asking questions. They also expressed that they felt at ease when talked with doctors talked who seemed friendly or approachable.

> *Sometimes doctors only have a certain amount of time, and I need to hurry up because they have other patients waiting for treatment, therefore, I couldn’t talk about much more things that bothered me…* (Participant 15)
>
> *Doctor gave me some confidence straight off, because he said ‘please let me know if there is anything you don’t understand or I haven’t explained it for you properly’, just a couple of sentences, but made me to be able to ask that question because I had the confidence to speak up…* (Participant 9)

#### 2.2.3. Motivation factors

##### 2.2.3.1. Reflective motivation

###### Outcome expectancies

The majority of participants expresses concerns about the safety and efficacy of anti-VEGF treatment, which affected their decisions on injection and led to missing the optimal opportunity.

> *It’s very hard to understand that I was diagnosed with DME and may be blindness without treatment, and my brain just went completely blank. So I told what doctor said to my family, and went to another hospital one month later after some hesitation, where I was diagnosed with DME again and doctor told me that I should receive anti-VEGF as soon as possible, but I was torn between injection and not injection due to the safety and effect…* (Participant 11)

##### 2.2.3.2. Automatic motivation

###### Fear of blindness

Fear of blindness made participants more cautious, and preferred to get advice from different doctors, especially in metropolis, which was one of the main obstacles to immediate treatment.

> *I was diagnosed with DME after several examinations and doctor told me I should receive intravitreal anti-VEGF injection to retard the progress, although I thought it was unbelievable, I accepted injection immediately due to fear of blindness …* (Participant 14)
>
> *When doctor told me that I should receive intravitreal anti-VEGF injection, I was afraid of the blindness and the risk of injection, so I went to another hospital after being diagnosed, I received the injection immediately…* (Participant 19)

###### Fear of injection

Fear of injection was another main obstacle to immediate treatment. Several participants pointed out that they experienced varying degree of fear before injection, some overcame their fears, others didn’t. As an alternative treatment, they chose eyedrops or oral drugs as their first choices.

> *I didn’t know how to inject medications into eyes before the doctor’s recommendation, and I thought it was unbelievable, so I was afraid of injection and also worried about it…* (Participant 22)
>
> *At that time, I was afraid of getting an injection and didn’t know what would happen if it hit my eyes. I still thought about conservative treatment, but later, conservative treatment failed, so I received anti-VEGF injection…* (Participant 18)
>
> *As the saying goes, there is no sand in your eyes, and if you want to get an injection into your eyes, it’s terrifying…* (Participant 5)

## 3. Discussion

This study provides a comprehensive understanding of patients’ perspectives on factors influencing their capability, opportunity and motivation to receive anti-VEGF treatment. Eleven factors, mostly commonly raised by participants, emerged from the COM-B domains, are included in this discussion

### 3.1. Capability factors

Lack of knowledge and high treatment expenditure were major barriers to timely receiving anti-VEGF treatment, categorized into the ‘capability’ theme. The majority of participants in our study were retirees with junior high school educational level, which led to the lack of sufficient knowledge of disease and influenced their decision on treatment. Thus, when something wrong with their eyes, they thought it was the normal aging process or ‘with cataract’. Disease knowledge play an important role in seeking treatment and changing behavior[12,13]. Lack of knowledge adversely affected the patient’s outcomes[14], and can be improved by providing disease knowledge education which can be added to pre-injection counseling. Meanwhile, patient’s knowledge about disease and medication, *and patient expectations about the treatment outcome* may also influence their adherence and persistence[15,16].

Some participants even thought ‘prioritized glycemic control rather than anti-VEGF’. This result might be attributed to the follow reasons: (1) Diagnosed with DME was earlier than diabetes, in other words, until the patient was diagnosed with DME, he found that he had diabetes. (2) Lack of knowledge of fundus disease. In China, public awareness of cataract is much higher than other eye diseases due to the popularization of cataract surgery and policy support. This is why participants always think that they had ‘cataract’.

Although medical insurance paid for partial treatment costs, some participants still felt that the remaining were too expensive to afford, which was another major obstacle to timely treatment, which was consistent with other studies[17], only 15% of the out-of-pocket expenditure patients reported high adherence to medication[18].

### 3.2. Opportunity factors

In our study, we found that some physical opportunities might lead to delayed diagnosis, untimely treatment and follow-up.

Some participants mentioned anti-VEGF injection was not available in their hometown and chose alternative treatment. Until symptoms were more severe than before, they finally referred to other hospitals to receive treatment. Lack of access to health-care services decreased adherence[16]. In China, intravitreal anti-VEGF injection was only available in tertiary hospitals where the costs can be paid by Chinese medical insurance.

‘Recommend eyedrops, laser and oral drug but not mentioned intravitreal anti-VEGF injection’, ‘as an optional therapy, selected more convenient treatment rather than anti-VEGF agents’ and ‘no confidence in treatment from doctor’ were all the obstacles to timely treatment. Some participants had to refer to other hospitals, after making sure anti-VEGF treatment was the optimal one, they received injection immediately. All of them expressed the need for doctor’s authoritative advice on intravitreal anti-VEGF injection and detailed introduction to the injection process which can help them receive standard treatment earlier. Individuals’ preferences are strongly affected by the way in which choices are presented[19].Some participants mentioned that it was difficult to communicate with doctors. Communication was identified as one of the most significant obstacles in terms of the subtheme “physical opportunity”[20]. Most patients in China went directly to tertiary hospitals regardless of the severity or type of disease, leading to an excessive clinical workload for doctors in tertiary hospitals[21], the accumulation of patients also leads to long waiting time and short reception time, and make doctors have no much time to give detailed explanation, the feeling of rushed by doctors made it difficult for patients to raise all their concerns or ask questions, which was consistent with other research where patients expressed a desire for longer consultations with their clinicians[22,23]. Other studies have found that multiple visits with a clinician can generate trust and comfort, and long-term relationships can promote patient participation[24]. Intravitreal anti-VEGF injection in patients with DME was not a one-time treatment, and needed a long-time regular follow-up and treatment, thus, effective communication and comfortable relationship play a crucial role[21].

### 3.3. Motivation factors

In terms of reflective motivation educed the subthemes ‘outcome expectancy’. Participants worried about the deterioration of vision acuity impacted on their daily life, which were the primary incentive to anti-VEGF treatment. Higher outcome expectancy was associated with higher self-management[25], adherence[26,27] and lower depression[27]. Therefore, we should attach importance to patient’s outcome expectancy and relations between it and adherence.

We also discovered two subthemes ‘fear of blindness’ and ‘fear of injection’ were partial causes for untimely treatment. ‘Fear of blindness’ made participants willing to injection, but meanwhile, ‘fear of injection’ made the refusing to injection. Therefore, timely alleviation of anxiety and fear of injection are crucial to the success of treatment, which requires a combination of assessment, education and behavioral intervention[28,29].

### 3.4. Limitation

This study only recruited patients with DME who received intravitreal anti-VEGF injection, and other clinical variables such as the severity and duration of disease, as well as other treatments patients, were not included. Despite these potential limitations, the strengths of our study included using the COM-B framework to guide the development of interview and analysis the results, enabling us to understand the factors affect patient’s behavior of intravitreal anti-VEGF injection, and providing evidence for optimizing the process of pre-injection counseling in order to accelerate decision-making and improve treatment adherence.

## 4. Conclusion

We identified eleven factors affected patient intravitreal anti-VEGF injection arising from interviews based on the COM-B model. The main obstacles to receiving timely intravitreal anti-VEGF injection are changeable, and effective interaction between clinicians and patients, as well as authoritative advice from clinicians, can promote patients to receive timely injection. Therefore, pre-injection consultation, care, and decision-making support can be based on these results.

## Data Availability

All data produced in the present work are contained in the manuscript

## Acknowledgements

Special thanks to the participants who volunteered to take part in this research.

## Contributors

LS and FQ were responsible for the conception and design of the study, compiling the interview schedule. LS, XY and PJN analyzed and interpreted data. LS, PJN, XX, TSY and DZY contributed substantially to conception and design, acquisition of data, and analysis and interpretation of data. LS and PJN drafted the article. FQ, XX, and TSY revised the article critically for important intellectual content. All authors have read and approved the final manuscript. All authors had full access to all of the data in the study and can take responsibility for the integrity of the data and the accuracy of the data analysis. LS is responsible for the overall content of the manuscript and serves as the guarantor.

## Patient and public involvement

Patients and/or the public were not involved in the design, or conduct, or reporting, or dissemination plans of this research.

## Patient consent for publication

Not applicable.

## Data availability statement

All data relevant to the study are included in thearticle.

